# Plasma Proteomic Signature of Frailty in 50,506 Adults

**DOI:** 10.1101/2025.06.13.25329572

**Authors:** Xueqing Jia, Weijing Gao, Jingyun Zhang, Yanjie Zhao, Liming Zhang, Xingqi Cao, Youheng Wu, Lina Ma, Liangkai Chen, Liang Sun, Huan Guo, Cuntai Zhang, Juulia Jylhävä, Zixin Hu, Hampus Hagelin, Sara Hägg, Emiel O. Hoogendijk, Zuyun Liu

**Author notes:** **Corresponding author:**Zuyun Liu, Professor, Center for Clinical Big Data and Analytics Second Affiliated Hospital, Department of Big Data in Health Science School of Public Health, Zhejiang Key Laboratory of Intelligent Preventive Medicine, Zhejiang University School of Medicine, Hangzhou 310058, Zhejiang, China. or; Zixin Hu, Associate Researcher, Artificial Intelligence Innovation and Incubation Institute, Fudan University, Shanghai 201203, China. Contributed equally to this work (Xueqing Jia, Weijing Gao).

## Abstract

Proteomics enables systematic elucidation of the biological mechanisms underlying health states including frailty. Here, through a large-scale proteome-wide association study (PWAS) encompassing 2,911 plasma proteins in 50,506 UK Biobank participants, we identified 1,339 proteins significantly associated with frailty, revealing novel functional modules implicated in frailty pathogenesis, particularly the one characterized by the collagen-containing extracellular matrix and vesicle lumen pathways. Replication analyses in an independent external cohort (TwinGene study) confirmed partial but consistent associations at both protein and pathway levels, supporting the reliability of these findings. Mendelian randomization analyses supported causal associations of 50 proteins with frailty. Protein-protein interaction network and expression quantitative trait loci analyses revealed MMP1 and LGALS8 serving as hub proteins. Moreover, we developed a novel proteome-based frailty measure, termed as Proteomic Frailty Score (PFS), which demonstrated robust predictive performance (C-index > 0.7) for 198 (30.2% = 198/655) incident diseases across 13 categories and broad responsiveness to 85 modifiable risk factors. Incorporating PFS into a conventional risk factors model significantly improved the predictive performance for 510 (77.9% = 510/655) incident diseases. Longitudinal analyses with three assessments (n∼1000) revealed an accelerated progression of the PFS with advancing age and increasing baseline frailty severity. To facilitate public use, we further created a publicly accessible online tool for PFS calculation (https://zipoa.shinyapps.io/frailty/). Finally, we observed a biphasic pattern of frailty-associated proteomic dysregulation across lifespan, with peak transitions occurring at approximately ages 50 and 63, implicating distinct biological pathways. Together, we establish PFS as a robust biomarker of biological aging while identifying critical windows and molecular targets for interventions against frailty progression.

## 1. Introduction

Frailty is an escalating global public health challenge, characterized by multisystem physiological decline and increased vulnerability to stressors, driving adverse health outcomes and imposing substantial healthcare burdens worldwide^1,2^. Despite decades of research, no globally standardized frailty assessment tool exists^2^. Current approaches, including the widely adopted frailty phenotype^3^ and frailty index (FI)^4^, predominantly rely on questionnaires, performance-based tests, or routine clinical data. These methods typically require specialized equipment and trained personnel for accurate administration, while also exhibiting limitations including inconsistent validation, restricted scalability and uncertain capacity to track frailty progression over time^2^, underscoring a critical gap in clinical applicability. Moreover, although efforts have been directed toward biomarker discovery for frailty (e.g., inflammatory indicators, oxidative stress markers, and metabolic profiles)^5–7^, the molecular mechanisms driving frailty remain poorly elucidated, impeding development of preventive programs and targeted intervention in the context of rapid population aging.

Proteomics offers a remarkable opportunity to systematically interrogate the protein profiles of health states, facilitating mechanistic understanding and biomarker identification, providing unprecedented insights to inform preventive strategies and advance personalized interventions^8,9^. However, existing proteomic studies on frailty often have small sample sizes (typically in hundreds to low thousands) or limited proteome coverage^10–12^. Furthermore, observational study designs are inherently limited by residual confounding and reverse causality, complicating the distinction between causal proteins and merely frailty-associated byproducts. Expanding proteome-wide association studies (PWAS) to larger cohorts, integrated with causal inference methodologies, holds promise for advancing our understanding of the biological basis of frailty. Moreover, while previous studies have revealed peripheral proteins associated with aging, the dynamic changes in frailty-specific plasma proteomes during aging remain unknown. Bridging this knowledge gap may pinpoint critical frailty progression phases, enabling timely preventive strategies and mechanistically targeted interventions.

Here, leveraging large-scale proteomic data from the UK Biobank (UKB), we first characterized the proteomic signatures of frailty (manipulated by FI) and elucidated its in-depth biological mechanisms. We then estimated the potential causal relationships between identified proteins and frailty through Mendelian randomization (MR) analyses. Next, we developed a novel proteome-based frailty assessment tool, termed as the “Proteomic Frailty Score (PFS)”, and evaluated its predictive performance for 655 incident diseases across 13 categories, as well as its responsiveness to 99 modifiable risk factors across six categories. To facilitate clinical utility, we further created a publicly accessible online tool for PFS calculation (https://zipoa.shinyapps.io/frailty/). Finally, applying differential expression-sliding window analysis (DE-SWAN), we identified two critical transition points in the frailty-associated proteome during aging. We validated our PWAS findings in an independent dataset (i.e., the TwinGene study), observing partially consistent associations at both protein and pathway levels that support the robustness of our results.

## 2. Results

### 2.1 Participants characteristics

Our primary study population included 50,506 participants from the UKB, with a median age of 58.6 years (interquartile ranges [IQR]: 50.7-64.0), of whom 53.9% were female and 94.1% were white ethnicity. All participants had quality-controlled proteomic data, > 39 FI items data and all covariates. A flow chart of the participant selection is shown in ***Figure S1***. Among them, 5,670 (11.2%) were identified as experiencing frailty. Detailed demographic characteristics in overall participants and by frailty status are provided in ***Table S1***.

### 2.2 Plasma proteome-wide association of frailty

To identify proteins associated with frailty, we conducted PWAS using linear regression on 2,911 plasma proteins, with FI (as continuous variable) as the outcome. After adjusting for multiple covariates, 1339 proteins showed significant associations with 1-standard deviation (SD) FI increase (***Figure 2a*** and ***Table S2***) at the Bonferroni-corrected threshold (*P* < 0.05/2911 = 1.7□×□10^−5^). Notably, 80.3% of these identified proteins exhibited positive associations, suggesting that elevated protein abundance may contribute to or reflect frailty pathophysiology (***Figure S2a)***. Similar results were observed when frailty was treated as a dichotomous variable (non-frail: FI ≤ 0.21 vs. frail: FI > 0.21^13^; ***Table S3*** and ***Figures S2b-c***). The proteomic associative patterns remained robust in analyses specifically performed in: 1) participants with complete data on all FI items (N = 41,821; ***Table S4*** and ***Figure S2d***), 2) the randomly selected subset that remains highly representative of the overall UKB participants^14^ (N = 43,455; ***Table S5*** and ***Figure S2e***), and 3) the Caucasian subgroup (N = 42,406; ***Table S6*** and ***Figure S2f***). These findings showed high correlations with those from the full sample (all r > 0.9, *P* < 2.2□×□10^−16^; ***Figures S2g-i***). Linear models incorporating interaction terms between age or sex and proteins associated with FI revealed significant interactions for 167 proteins with age and 16 proteins with sex (*P* < 0.05/1339 = 3.7×10^−5^; ***Table S7***). Detailed PWAS results stratified by age groups (younger: < 60 years, N = 27,867; older: ≥ 60 years, N = 22,639) and sex (males: N = 23,262; females: N = 27,244) are illustrated in ***Tables S8-9*** and ***Figures S3a-f***.

To further elucidate the biological functions of proteins associated with frailty, we investigated their involvement in functional pathways. Following false discovery rate (FDR) correction, several immune-related pathways emerged as the most significantly enriched, such as leukocyte migration, cytokine-cytokine receptor interactions, and neutrophil degranulation (***Figure 2b*** and ***Table S10***). These findings align with prior studies emphasizing immune dysregulation in frailty pathophysiology^15^. Using the approach proposed by Shen et al. (***Methods***)^16^, we further eliminated redundant pathways and delineated distinct functional modules associated with frailty (***Tables S11-12***). Beyond previously reported modules, such as those related to immune-inflammatory processes^11,12^, we uncovered several novel functional modules. For instance, we identified a module characterized by collagen-containing extracellular matrix (adjusted *P* = 1.64□×□10^−6^) and vesicle lumen (adjusted *P =* 6.36□×□10^−7^) (***Figure 2c***). The collagen-containing extracellular matrix is pivotal for maintaining tissue structural integrity and stability^17,18^. Age-associated collagen degradation could undermine tissue resilience^19,20^, potentially driving the physical manifestations of frailty. Similarly, the vesicle lumen, crucial for cellular transport and signaling, exhibits functional decline with aging, impairing repair mechanisms and compromising physiological homeostasis^21,22^. Additionally, both pathways are closely linked to immune-senescence and inflammaging^22–24^, suggesting their indirect contributions to frailty pathogenesis. Together, these insights provide a more comprehensive understanding of the biological basis of frailty and highlight potential targets for future interventions.

To elucidate organ-specific expression patterns of proteins associated with frailty, we further conducted an enrichment analysis using the putative organ-specific plasma proteome encompassing 18 organ systems^25^ (***Methods***). Among the 1339 frailty-associated proteins identified above, only 323 (24.1%) were classified as organ-specific, directly supporting the notion that frailty is a systemic condition characterized by multisystem dysregulation^1^. These 323 proteins exhibited significant enrichment in 10 organs (false discovery rate [FDR] < 0.05), particularly in the liver and gastrointestinal system (e.g., stomach, intestine, pancreas), whereas no significant enrichment was observed in tissues such as heart, immune system, or kidneys (***Figure 2d*** and ***Table S13***). These findings suggest that certain organs may play distinct roles in the pathophysiology of frailty. Specifically, the pronounced enrichment observed in the liver and gastrointestinal system may reflect their central roles in metabolic regulation, detoxification, and nutrient absorption during frailty progression.

### 2.3 Replication of PWAS in the TwinGene cohort

To validate our PWAS findings, we replicated the analysis in an independent external cohort—the TwinGene study—which included 5,446 participants (mean age, 64.33 years) with serum samples (***Methods; Table S14***). Of the 1339 frailty-associated proteins identified in the UKB, only 297 were measured in the TwinGene cohort and were thus available for replication. Despite this limited overlap, we observed similar associations between these 297 proteins and FI across both cohorts (Pearson’s *r*□=□0.72; ***Figure 2e***). Among these proteins, 142 (47.8%) exhibited nominal significance, while 109 (36.7%) remained significant after FDR correction (*FDR < 0.05*), with 108 showing directionally consistent effects compared to those in the UKB (***Table S15)***. These results may be partly attributed to the relatively small sample size of the TwinGene cohort, differences in participant characteristics (TwinGene participants were on average over five years older than those in the UKB), as well as variations in sample type (serum vs. plasma) and assay methodology (***Methods***).

Furthermore, we investigated the functional pathways enriched among these replicated proteins and found that several key pathways previously highlighted in the UKB analysis, such as collagen-containing extracellular matrix and cytokine-cytokine receptor interactions, remained significantly enriched (***Figure 2f and Table S16)***. This cross-cohort consistency supports the robustness of our initial pathway-level findings and further highlights the biological relevance of these processes in the pathophysiology of frailty.

### 2.4 MR analysis between plasma proteins and frailty

To identify proteins exerting potential causal effects on frailty, we performed proteome-wide two-sample MR analyses for significant protein-frailty associations utilizing protein quantitative trait loci (pQTL) data^26^ and genome-wide association study (GWAS) summary statistics for FI^27^. Among the 1,339 significant proteins identified in the PWAS, 715 with available instrumental variables (IVs) were analyzed (***Methods***). Using the Wald ratio or inverse-variance weighted (IVW) method, we identified 50 proteins nominally associated with frailty (*P* < 0.05; ***Figure 2g and Tables S17-20***). To explore functional interactions among these candidate proteins, we constructed the protein-protein interaction (PPI) networks using the STRING database, identifying ten hub proteins (i.e., CDH1, ACE, MMP1, NCAM1, B2M, MMP3, PON1, MET, SORT1, LGALS8) via the maximal clique centrality (MCC) method (***Figure S4a)***.

To further validate the potential causal effects of the ten hub proteins at the gene level, we performed MR analyses using cis-eQTL data from the eQTLGen consortium^28^. Seven genes were analyzed (*MMP3*, *PON1*, and *MET* were excluded due to unavailable genetic data), among which *MMP1* and *LGALS8* exhibited significant causal associations with frailty, with effect directions consistent with their protein-level associations (***Table S21)***. We then performed phenome-wide MR (PheMR) analyses to examine associations between MMP1/LGALS8 and 2,803 phenotypes using data from the FinnGen study^29^. *MMP1* showed significant causal associations with 127 phenotypes, including positive associations with 78 phenotypes (e.g., chronic hepatitis and factitial dermatitis) (***Table S22)***. *LGALS8* exhibited significant causal associations with 229 phenotypes, including negative associations with 197 phenotypes (e.g., asthma and stroke) (***Table S23)***. Focusing on aging-related diseases within these significant associations (n = 18 out of 78 for MMP1; n = 22 out of 197 for LGALS8), we implemented colocalization analyses to address potential linkage disequilibrium (LD) confounding. These analyses revealed strong colocalization evidence for MMP1 with chronic obstructive pulmonary disease (COPD; PPH3 + PPH4 > 0.8), as well as colocalization evidence for LGALS8 with type 2 diabetes (PPH3 + PPH4 > 0.8) and heart failure (PPH3 + PPH4 > 0.7) (***Table S24 and Figure S8)***. The findings suggest potential causal roles of MMP1 and LGALS8 in frailty and age-related diseases, although further investigation into the mechanisms and the roles of other hub proteins is needed.

By leveraging single-cell RNA sequencing (scRNA-seq) data of peripheral blood mononuclear cell (PBMC) from both healthy and frail older participants^30^, we profiled the cellular expression levels of *MMP1* and *LGALS8*. We found that *LGALS8* was relatively highly expressed in monocytes, T cells, and NK cells, whereas *MMP1* showed low expression across all blood cell types examined (***Figure S4b***). Notably, differential expression analyses revealed that *LGALS8* was significantly downregulated in monocytes and NK cells of frail participants (*P* < 0.05) (***Figure S4c***).

### 2.5 PFS outperforms in predicting health-related outcomes

Using the 1,339 FI-associated proteins identified through PWAS, we developed PFS by applying a ten-fold iterative scheme of the least absolute shrinkage and selection operator (LASSO) algorithm with ten-fold cross-validation (***Figure 1 and Methods***). This approach generated ten sparse models and produced one hold-out set-derived prediction for each participant. The resulting models retained 432 to 512 proteins across folds (***Tables S25-27***). PFS showed moderate predictive performance for FI, with a mean out-of-sample correlation coefficient of 0.503 (***Figure 3a and Table S28*)**. Consistent with FI (***Figure S5***), PFS was significantly elevated in males (vs. females) and older adults (vs. younger) (both *P* < 0.001) (***Figure 3b***). To facilitate public use of PFS, we provided an illustrative online tool (https://zipoa.shinyapps.io/frailty/) (***Figure S6***).

**Figure 1.**
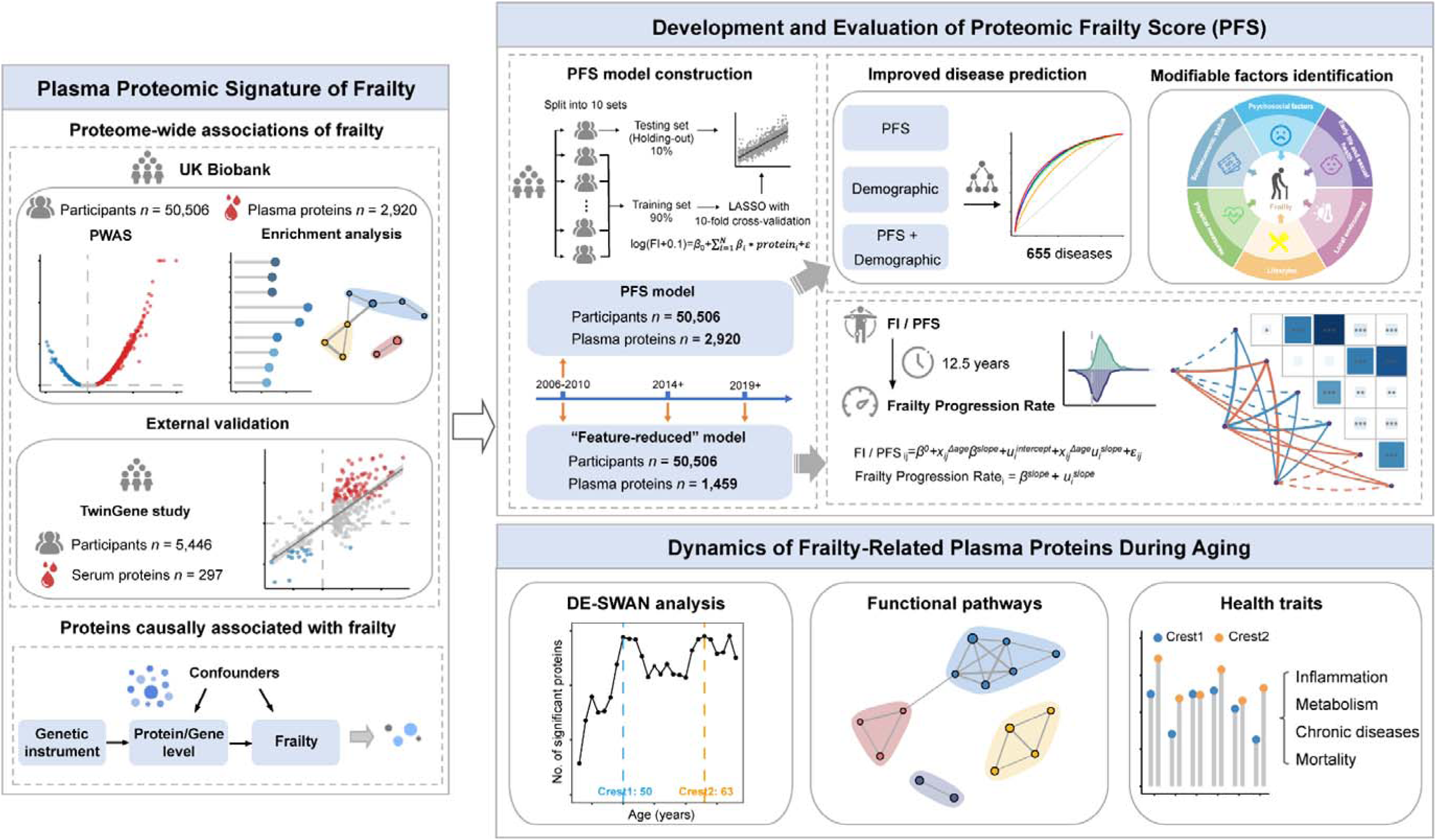
The roadmap of this study. Left part, we used the plasma proteomic data of 50,506 UK Biobank (UKB) participants to conduct proteome-wide association analysis (PWAS) of frailty, and further elucidate the biological functions of proteins associated with frailty. We then validated our PWAS findings in an independent external cohort with 5,446 participants (i.e., the TwinGene study). Next, we performed a proteome-wide two-sample Mendelian randomization (MR) analysis for significant protein-frailty associations to identify proteins with causal roles in frailty, utilizing protein quantitative trait loci data and genome-wide association study summary statistics for frailty. Top right part, the development and evaluation of proteomic frailty index (PFS) model. First, using the 1,339 frailty-associated proteins identified through PWAS, we developed PFS by applying a ten-fold iterative scheme of the least absolute shrinkage and selection operator algorithm with ten-fold cross-validation. Then we evaluated the predictive performance of PFS for disease risk and compared it with conventional factors-based model, as well as their integration. We investigated the associations of 99 potentially modifiable factors across six categories from the UKB baseline survey with PFS. For longitudinal analyses, we focused on 1,459 proteins with less than 20% missing values across all three visits (baseline, third, and fourth) and retrained the PFS model (“feature-reduced” models) in the 50,506 participants at baseline using these proteins. We then estimated the PFS (and FI) change rate for 984 individuals with quality-controlled proteomic data at all three visits. We then evaluated the associations of the PFS (and FI) change rate with baseline age, frailty severity, and accumulated diseases counts. Bottom right part, we performed a differential expression-sliding window analysis (DE-SWAN) to comprehensively characterize changes in FI-associated proteins across the human lifespan. Two prominent peaks occurring around ages 50 and 63 were identified. We then conducted separate functional enrichment analyses for proteins specific to each crest to further elucidate the biological functions associated with each peak. Finally, we tested the associations between proteins of the two crests and health-related traits.

**Figure 2.**
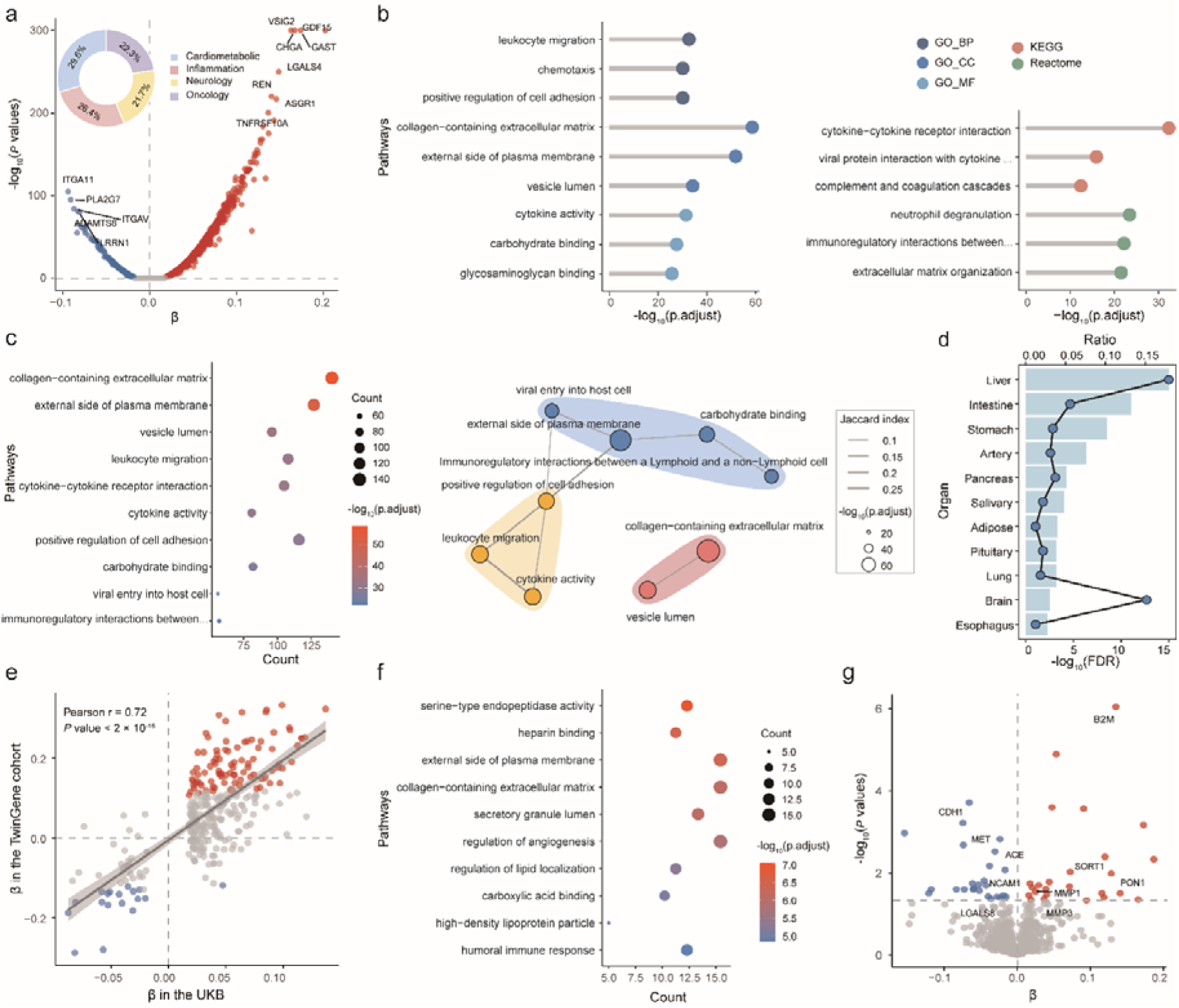
Plasma proteomic profiles of frailty. **a**. Volcano plot showing associations between 2,911 plasma proteins and frailty. The *x* axis represents β coefficients, and the *y* axis represents -log_10_(*P* values). The dashed lines indicate the thresholds for Bonferroni corrections (*P*<□0.05/2911=1.7□×□10^−5^). The pie charts depict the proportions of identified proteins across four distinct panels. The linear regression models were adjusted for chronological age, sex, the first 20 genetic principal components, ethnicity, educational level, Townsend deprivation index, alcohol consumption, smoking status, regular exercise, and body mass index. **b.** The top three enriched Gene Ontology terms and KEGG/Reactome pathways for frailty-associated proteins. BP, Biological Process; CC, Cellular Component, MF, Molecular Function. **c.** Functional module analysis for frailty-associated proteins. The left panel is the top ten representive pathways after eliminating redundant pathways. The right panel is the pathway similarity network. **d.** Enrichment of frailty-associated proteins across 18 organ systems. The bar plot indicates the -log_10_(FDR), reflecting the statistical significance of enrichment. *P* values were calculated using Fisher’s exact test. Each dot represents the ratio of frailty-associated proteins specific to an organ relative to the total number of organ-specific proteins. **e.** Scatter plot showing the correlation of β coefficients between 297 proteins and frailty in the UKB and the TwinGene cohorts. The coefficient and *P* values obtained from Pearson correlation analysis were shown in the left top part. Red dots represent proteins positively associated with frailty; blue dots represent negatively associated proteins. **f.** Top ten representive pathways for frailty-associated proteins in the TwinGene cohort after eliminating redundant pathways. **g.** Causal associations from proteome-wide mendelian randomization (PWMR) study using Wald ratio or inverse-variance weighted (IVW) methods. The *x* axis represents β coefficients, and the *y* axis represents -log_10_(*P* values). The dashed lines indicate *P*<□0.05.

**Figure 3.**
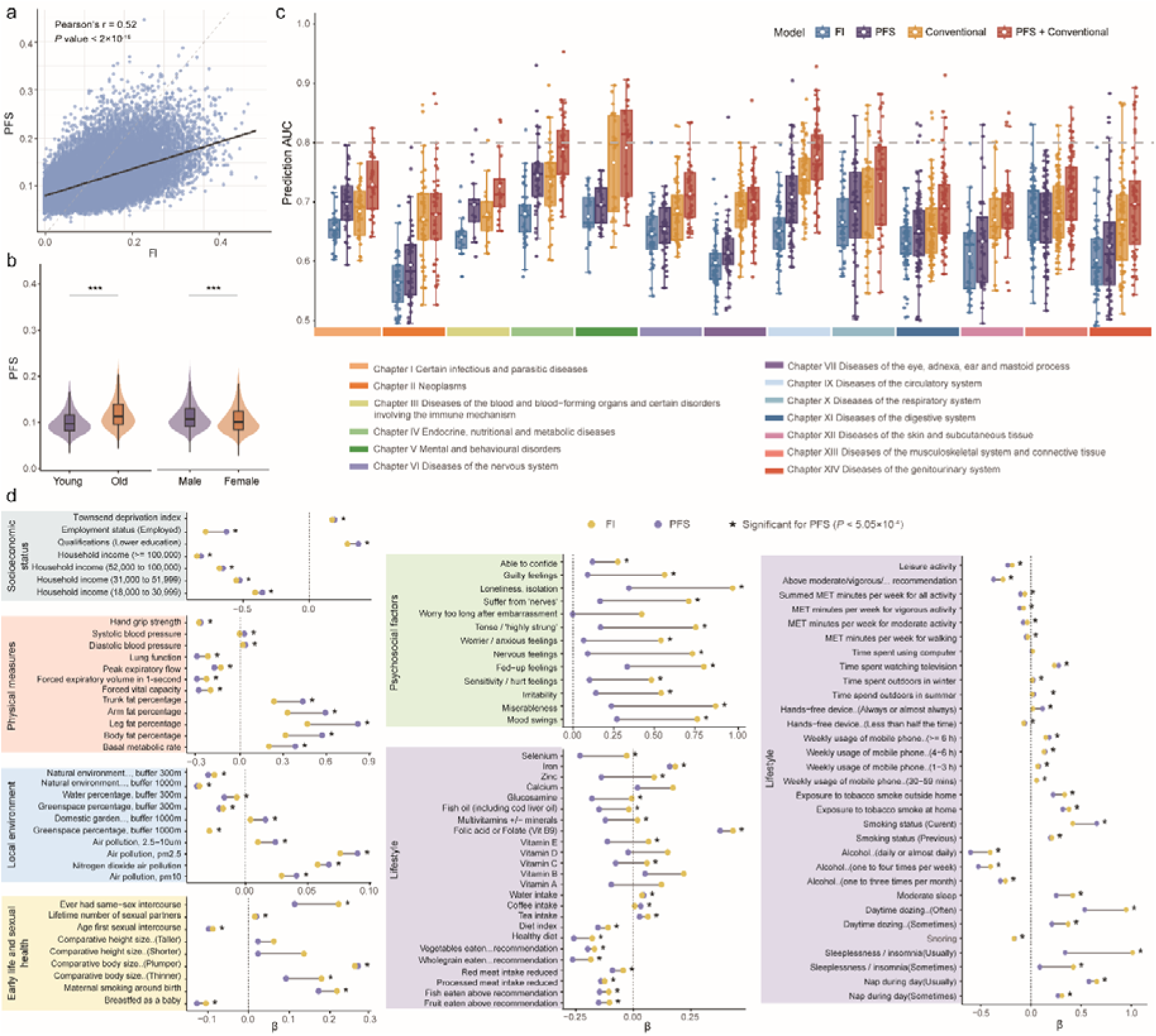
The Proteomic Frailty Score (PFS) improves disease prediction and responds to key modifiable risk factors. **a.** Scatter plot showing the correlation between PFS and traditional FI across all participants. Each scatter indicates a single participant. The Pearson’s correlation coefficient (r) is shown in the left top part. **b.** Violin plots comparing PFS distributions by sex (males: N=23,262; females: N=27,244) and age group (younger: <60 years, N=27,867; older: ≥60 years, N=22,639). ***indicates p<0.001 (two-tailed t-test). **c.** The discriminative performances for diseases prediction (quantified by area under the curve [AUC]) across four models: traditional FI, PFS, conventional risk factors model, and PFS + conventional risk factors model. The box plot shows the median (line), interquartile range (IQR) (box) and whiskers extending to 1.5 × IQR. Hollow circles represent mean values. The conventional risk factors included age, sex, ethnicity, Townsend deprivation index, body mass index, systolic blood pressure, and status of smoking and alcohol consumption. **d.** The associations of PFS and traditional FI with 99 modifiable risk factors across six categories. The linear models were adjusted for age, sex, and ethnicity. Only factors significantly associated with PFS or FI (*P* < 0.05/99 = 5.051710^−4^) were included in the plot. Asterisks indicate factors significantly associated with PFS.

We then evaluated the predictive performance of PFS for health-related outcomes (i.e., all-cause mortality, cause-specific mortality, and 655 incident diseases), and compared it with conventional factors-based models, as well as their integration (***Figure 3c and Tables S29-33*)**. PFS demonstrated strong predictive capability for all-cause mortality (C-index = 0.708) and cause-specific mortality (e.g., cardiovascular disease [CVD]-related death: C-index = 0.763; respiratory disease-related death: C-index = 0.828). Additionally, PFS achieved a C-index exceeding 0.70 for 198 incident diseases (30.2% = 198/655) across 13 disease categories. Notably, PFS achieved excellent predictions (C-index > 0.8) for 29 diseases, particularly in endocrine, nutritional and metabolic (n = 7 out of 42), and circulatory (n = 8 out of 65) diseases, such as type 2 diabetes with peripheral circulatory complications (C-index = 0.930; 95%CI: 0.909, 0.951), hypertensive renal disease (C-index = 0.905; 95%CI: 0.885, 0.924), and chronic nephritic syndrome (C-index = 0.883; 95%CI: 0.855, 0.911) (***Table S32)***. PFS significantly outperformed conventional factors-based model in predicting 122 diseases (18.6% = 122/655), such as diabetic neuropathy (PFS: C-index = 0.708 vs. conventional factors model: C-index = 0.603, *P* < 0.05). Furthermore, integrating PFS with conventional factors significantly improved predictive accuracy for 510 diseases (77.9% = 510/655) (***Table S33)***.

PFS also demonstrated significantly better predictive performance than FI for 299 incident diseases (45.6% = 299/655) across 13 disease categories, particularly in circulatory (n = 56 out of 65) and endocrine, nutritional, and metabolic (n = 36 out of 42) diseases. When integrated with conventional factors, models incorporating PFS continued to outperform those incorporating FI for 178 diseases. In contrast, FI demonstrated significantly better predictive performance than PFS for 51 incident diseases (7.8% = 51/655) across seven disease categories, particularly in diseases of the musculoskeletal system and connective tissue (n = 23 out of 96), and mental and behavioral disorders (n = 5 out of 24) diseases. Furthermore, combining PFS with conventional factors and FI further improved predictive accuracy compared to models including only conventional factors and FI, with significant enhancements observed for 408 diseases (62.3% = 407/655) (***Table S33***). These findings suggest that PFS captures additional predictive information not encompassed by FI.

### 2.6 PFS broadly responds to modifiable risk factors

We further investigated the associations of 99 potentially modifiable risk factors from the UKB baseline survey with frailty, as measured by PFS and FI. After Bonferroni correction, 85 out of 99 risk factors were significantly associated with PFS (P < 5.05×10^−4^, ***Figure 3d* and *Table S34***). Among the top 20 factors, 10 were positively associated with frailty, such as leg fat percentage (β = 0.819; 95% CI:□ 0.804, 0.834), smoking status (current vs. never: β = 0.656; 95% CI:□ 0.629, 0.683), and Townsend deprivation index (TDI, β = 0.192; 95% CI:□0.184, 0.200); and 10 factors were negatively associated with frailty, such as current employment status (employed vs. unemployed: β = −0.627; 95% CI:□−0.657, −0.597), forced expiratory volume in 1-second (β = −0.3; 95% CI: −0.311, −0.289), and diet index (β = −0.154; 95% CI: −0.162, −0.146). Similar exposure-wide analysis pattern was observed for FI. Comparable findings were also noted when stratifying by age and sex (***Table S34***).

Interestingly, we found that the responses of PFS and FI to different categories of modifiable risk factors exhibited notable differences (***Figure 3d***). For instance, FI showed stronger associations with psychosocial factors (e.g., nervous feelings, worrier/anxious feelings, loneliness) and sleep-related measures (e.g., sleeplessness/insomnia), likely due to its reliance on self-reported psychological and functional status, making it more sensitive to subjective factors. In contrast, PFS showed stronger associations with physical measures (e.g., pulmonary function, body composition), dietary quality, and physical activity, which are closely linked to biological changes (e.g., inflammation, metabolic proteins). Thus, PFS, rooted in objective proteomic data, showed more sensitivity to capturing these biologically driven changes compared to FI.

### 2.7 Longitudinal PFS changes facilitate sensitive dynamic monitoring of frailty

A subset of ∼1,000 participants had proteomic data available at their third and fourth visits, enabling us to evaluate frailty progression based on changes in PFS and FI. For longitudinal analyses, we focused on 1,459 proteins with less than 20% missing values across all three visits (baseline, third, and fourth) and retrained the PFS model (“feature-reduced” models) in the 50,506 participants at baseline using these proteins (***Tables S35-37***). The feature-reduced model demonstrated strong correlation with FI (***Tables S38***). We then estimated the PFS (and FI) change rate for 784 participants with quality-controlled proteome and visit date data at all three visits (***Methods***). As expected, longitudinal analysis revealed frailty progression in most participants (90.8% by PFS; 70.8% by FI) (***Figure 4a***). Notably, frailty (evaluated by PFS) progressed more rapidly in older adults (≥ 60 years) compared to younger adults (< 60 years) (*P* < 0.001, ***Figure 4b***). Participants with accelerated frailty progression accumulated more diseases; each 1-SD increase in FI and PFS was associated with an average of 0.6 and 2.6 additional International Classification of Diseases, 10th edition (ICD-10) disease annotations, respectively (***Figures 4c-d and Table S39***). These findings underscore the importance of managing FI progression to mitigate disease burden.

**Figure 4.**
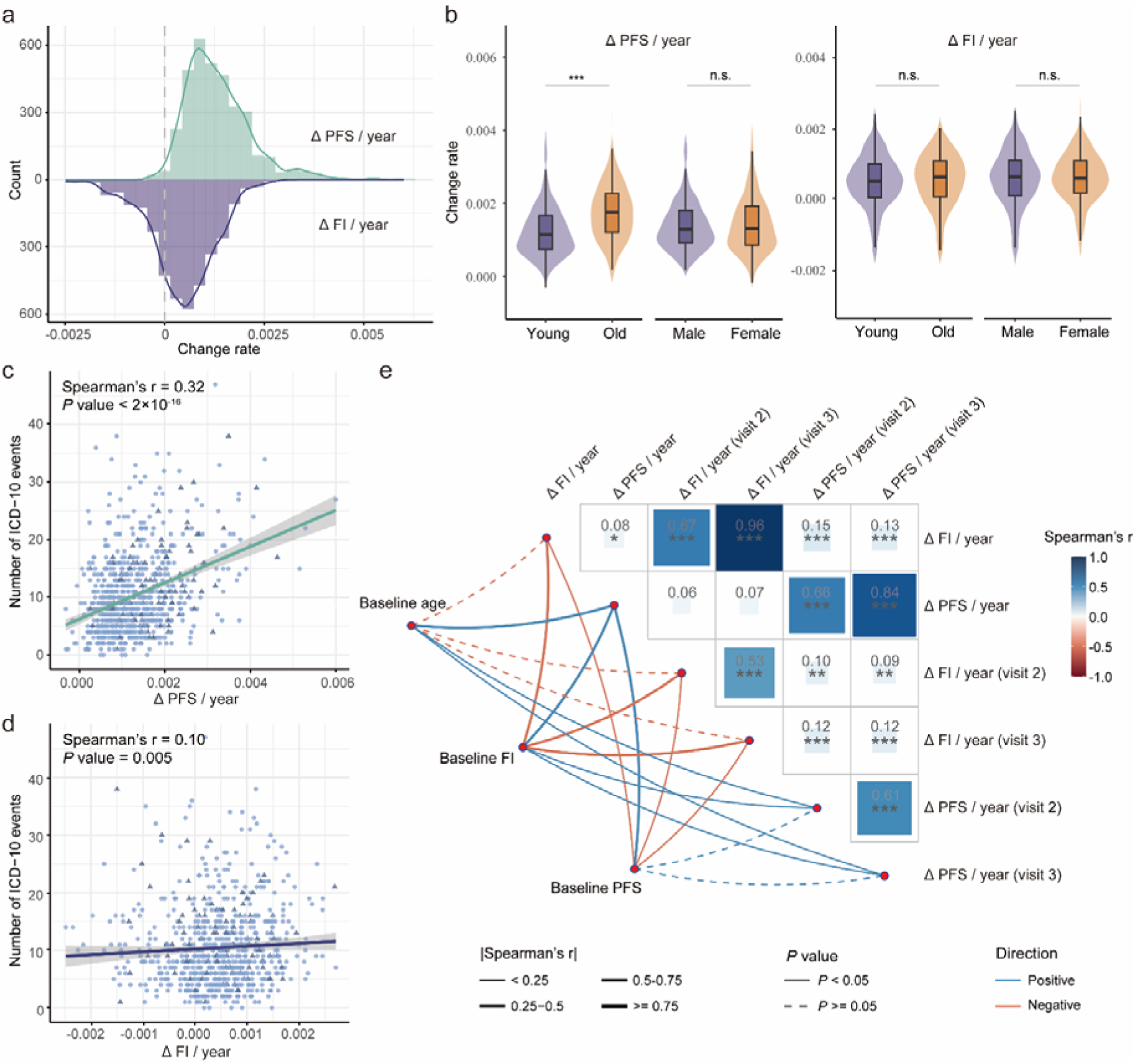
The distinct progression patterns between PFS and traditional FI during aging. **a.** Histogram comparing annualized progression rates (Δ/year) of PFS (derived from “feature-reduced” model) versus traditional FI. ΔPFS/year and ΔFI/year indicated the frailty progression rate calculated through mixed-effects model. **b.** Violin plots comparing the distributions of progression rates stratified by sex and age group (<60 vs. ≥60 years). ***p<0.001, two-tailed t-test; n.s., not significant (p ≥ 0.05). Boxplots within violins show median (center line) and interquartile range (IQR) (box bounds). **c, d.** Scatter plot showing the correlation of the change rates of PFS and traditional FI with accumulative disease count. The Spearman’s correlation coefficient (r) are shown in the left top part. **e.** The correlation heatmap showing the correlations of baseline age and frailty status (indicated by PFS and traditional FI) with frailty progression pattern (quantified by the change rate of PFS and traditional FI). ΔPFS/year(visit2) and ΔFI/year(visit2) indicated the frailty progression rate calculated by subtracting the baseline round from the third round and dividing by the follow-up time. ΔPFS/year(visit3) and ΔFI/year(visit3) indicated the frailty progression rate calculated by subtracting the baseline round from the fourth round and dividing by the follow-up time.

We further investigated how initial frailty status impacts progression rates. Interestingly, we found that baseline PFS and FI were negatively associated with the change rate of FI but positively associated with the change rate of PFS (***Figure 4e***). This suggests that FI may be less sensitive in capturing frailty progression, particularly among individuals with advanced aging and severe frailty, whereas the PFS demonstrates unique capacity for dynamically monitoring frailty even in this vulnerable population. These findings remained consistent when using an alternative method to calculate frailty progression rates (subtracting the previous round from the subsequent round and dividing by the follow-up time) (***Figure 4e***).

Next, we defined participants with FI change rates in the highest quintile as accelerated frailty progressors, and those in the remaining quintiles as normal progressors. Using the same threshold derived from the FI change rates, participants were categorized based on their PFS change rates. Participants were then classified into matched or mismatched groups according to whether their FI- and PFS-based progression status agreed. Notably, we found that mismatched individuals in the normal progressor group (i.e., accelerated progressors defined by PFS change rates) accumulated significantly more diseases, while those in the accelerated progressor group (i.e., normal progressors defined by PFS change rates) accumulated significantly fewer (***Figure S7***). This pattern echoes our hypothesis that PFS may be more sensitive to capturing early frailty progression.

### 2.8 Biphasic patterns of frailty-associated proteomic dysregulation across lifespan

Given the undulating nature of the aging plasma proteome, we performed a DE-SWAN analysis to comprehensively characterize changes in frailty-associated proteins across the human lifespan (***Methods***). We identified two prominent crests occurring around ages 50 and 63, which remained robust across different q-value thresholds (***Figures 5a-b***). The proteins associated with these two crests are detailed in ***Table S40***.

**Figure 5.**
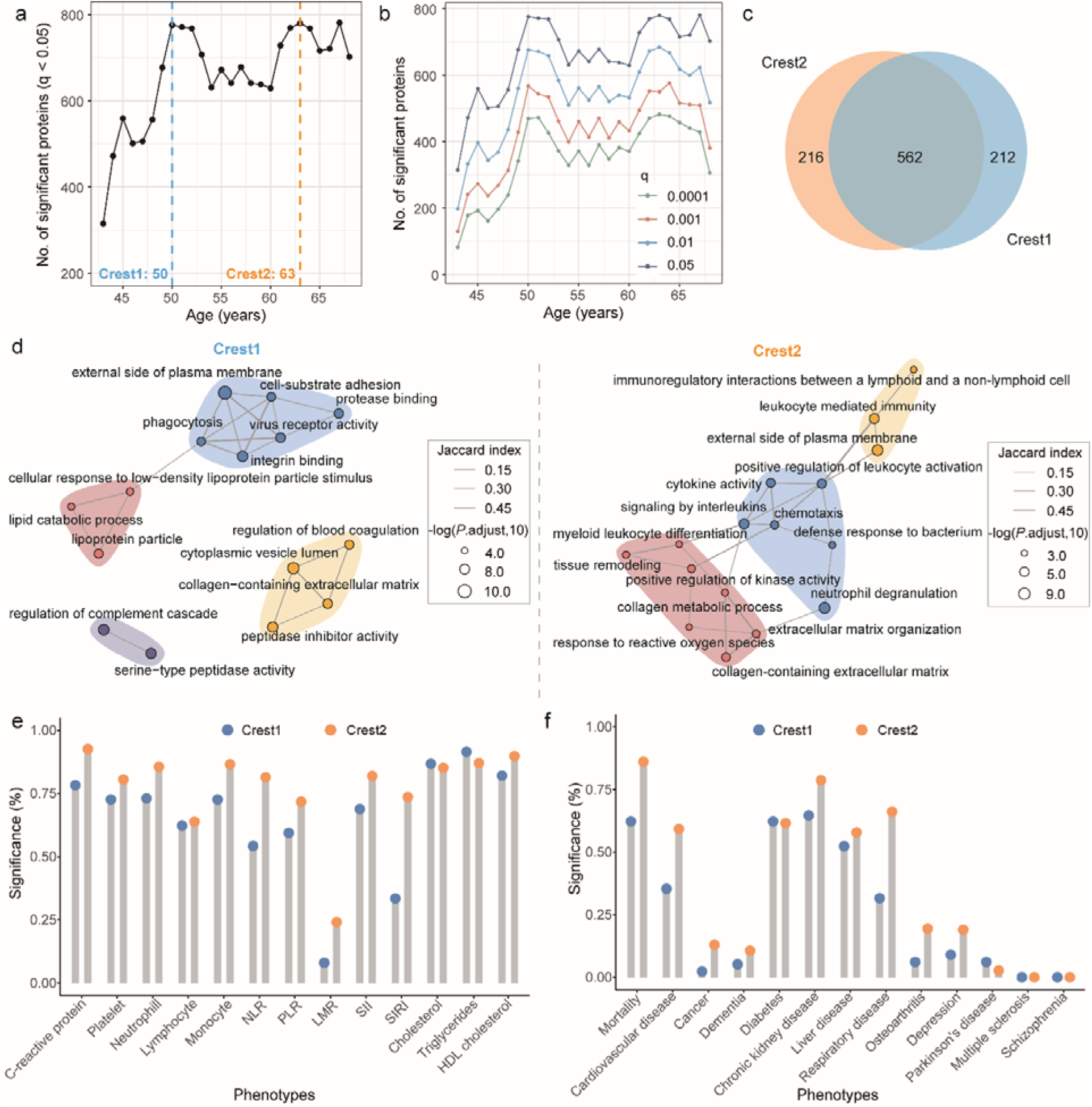
Biphasic waves of frailty-associated proteomic changes during aging. **a**. Age-dependent changes of frailty-associated proteins, showing two distinct peaks at 50 and 63 years. The y-axis displays counts of differentially expressed proteins. **b.** Robustness analysis demonstrating consistent detection of proteomic waves across multiple significance thresholds (*q*<0.0001, 0.001, 0.01, and 0.05). **c.** Venn diagram quantifying overlap between proteins identified in the 50-year (early) and 63-year (late) peaks. **d.** Pathway enrichment and module analysis for proteins specific to each crest. **d, e.** Barplots showing the proportions of significant associations between proteins of each crest and metabolic, inflammatory indicators (d) and age-related diseases (e). Associations with metabolic/inflammatory markers were assessed using linear regression, while age-related disease associations were evaluated using Cox proportional hazards models. All models were adjusted for chronological age, sex, the first 20 genetic principal components, ethnicity, educational level, Townsend deprivation index, alcohol consumption, smoking status, regular exercise, and body mass index.

To further elucidate the biological functions associated with each crest, we conducted separate functional enrichment analyses for proteins specific to each crest (***Figure 5c***). Proteins at both crests were enriched in several common pathways, such as external side of the plasma membrane and collagen-containing extracellular matrix organization, highlighting the critical role of cellular adhesion and structural integrity in aging and frailty development. However, each crest also exhibited distinct pathway enrichments, reflecting stage-specific biological processes. At age 50, proteins were particularly enriched in metabolic and signaling pathways, such as cholesterol metabolism, lipid catabolic process, carbohydrate derivative catabolic process, and hormone activity. Whereas proteins associated with the peak at age 63 were particularly enriched in pathways related to immune regulation (e.g., leukocyte mediated immunity), cytokine activity (e.g., cytokine-cytokine receptor interaction), and defense responses to pathogens (e.g., defense response to bacterium) (***Figure 5d*** and ***Tables S41-43*)**. These findings indicated that frailty progression with aging is a dynamic, undulating process characterized by waves of changes in plasma proteomes that reflect alterations in underlying biological processes.

We further tested associations between the proteins of two key crests and health-related traits including diseases and metabolic and inflammatory indicators. We found that proteins at age 50□years showed most associations with metabolic health traits (e.g., total cholesterol [TC], triglycerides [TG], and diabetes) and Parkinson’s disease, while proteins at age 63□years showed most associations with multisystem diseases (e.g., dementia, CVD, and cancer) and immune-inflammatory markers (***Figure 5e-f and Tables S44-45***).

## 3. Discussion

This large-scale proteomic analysis of 50,506 UKB participants identified 1,339 plasma proteins significantly associated with frailty, providing unprecedented insights into its proteomic landscape. Through comprehensive functional annotation and MR analyses, we elucidated novel biological pathways implicated in frailty pathogenesis and suggested potential causal roles of several candidate proteins in driving its progression. Replication analyses in the TwinGene cohort (n = 5,446) confirmed partial but consistent associations at both protein and pathway levels, supporting the reliability of the above findings. Furthermore, We developed a novel proteome-based measure of frailty—PFS, which exhibited robust predictive capacity for a wide spectrum of incident diseases and broad responsiveness to diverse modifiable risk factors, highlighting its potential as a versatile aging biomarker. Longitudinal analyses with three assessments (n∼1000) revealed an accelerated progression pattern of PFS with advancing age and baseline frailty severity. In addition, we uncovered dynamic changes in frailty-associated proteins across the lifespan, with proteomic alterations peaking in the 50 and 63 ages, pinpointing critical windows for targeted interventions to mitigate age-related decline.

Previous studies have sought to delineate proteomic biomarkers of frailty in 880 community-dwelling Ashkenazi Jewish older adults (≥ 65 years)^11^ and 3,838 older adults (≥ 65 years) from the Atherosclerosis Risk in Community (ARIC) study^12^. This study profiled the plasma proteomic landscape of frailty in the largest cohort spanning a wider age range to date. Furthermore, we employed a comprehensive MR framework to demonstrate causal relationships between specific proteins and frailty, strengthening the evidence for their biological relevance. Complemented by in-depth functional exploration, we confirmed several frailty-associated proteins (e.g., FABP4, FSTL3, IL1RN, and PXDNL) and functional pathways (e.g., immune-inflammatory processes) consistent with prior research^11,12^, while also identifying novel proteins and pathways potentially involved in frailty development. For example, our study demonstrates that MMP1 and LGALS8 serve as biomarkers of frailty at both protein and gene levels. MMP1, a key component of the senescence-associated secretory phenotype (SASP), plays crucial roles in several physiological processes including inflammation, skin aging, and tissue remodeling^32^. Growing evidence supports its involvement in various age-related pathologies, particularly skin disorders^33^, COPD^34^, and cancer progression^35^. LGALS8, also known as galectin-8 (Gal-8), is a glycan-binding protein that critically modulates diverse cellular processes including adhesion, proliferation, spreading, and apoptosis^36^. Dysregulated Gal-8 functions have been observed in rheumatic, autoimmune and inflammatory disorders^37,38^. In short, by integrating large-scale proteomic profiling with causal inference methods, our findings advance the understanding of frailty biology and highlight promising targets for further mechanistic and therapeutic exploration.

Our newly developed PFS demonstrated predictive ability for multiple incident diseases that was comparable to or even exceeded conventional risk factors, while also providing additional predictive value beyond these established factors. These findings underscore the potential of PFS serving both as a streamlined alternative for conventional risk assessments and as a complementary tool to enhance multi-disease risk stratification. Furthermore, the wide spectrum of diseases included in our study enabled us to reveal distinct predictive efficacy between the traditional FI and PFS across disease categories. Notably, PFS demonstrated superior performance for the majority of diseases, particularly circulatory diseases and endocrine, nutritional, and metabolic disorders. This highlights the inherent advantage of proteomics in capturing dynamic molecular changes^39^ and underscores the broader applicability of PFS as a robust tool for frailty assessment. In contrast, the traditional FI, which incorporates clinical and functional measures^4^, better captured chronic physical and psychological impairments associated with musculoskeletal and mental health conditions.

Frailty research has progressively identified risk factors across sociodemographic, clinical, lifestyle, and biological domains, with modifiable factors being of particular interest for interventions^40,41^. Our study advances this field by systematically evaluating 99 potentially modifiable risk factors across six categories. The broad responsiveness of PFS to diverse modifiable risk factors underscores its significant potential as a versatile biomarker of aging^42,43^, highlighting the importance of multidimensional interventions for frailty mitigation. Interestingly, while both PFS and traditional FI capture known frailty risk factors, they exhibit distinct exposome association patterns. PFS demonstrates particular sensitivity to factors (e.g., lifestyle and physical measures) that typically require extended periods to manifest phenotypically, suggesting its ability to identify molecular perturbations preceding clinical manifestations^44^. This preclinical detection capability enables more timely evaluation of intervention efficacy and optimization of therapeutic strategies, representing a significant advantage over traditional FI’s phenotype-dependent assessments that are subject to reporting biases and delayed manifestations. Conversely, the traditional FI demonstrates the significance for monitoring interventions targeting mental health and sleep quality, such as cognitive behavioral therapy and social support programs. Together with findings on disease prediction aforementioned, these divergent exposome patterns suggests the complementary potential of combining both FI measures for a more holistic approach for assessing frailty, predicting disease risk, and evaluating intervention effectiveness, ultimately enabling more personalized frailty prevention and management strategies.

Our longitudinal analysis reveals distinct progression patterns between PFS and traditional FI. The PFS exhibits an age- and baseline frailty severity-dependent acceleration in progression rate, aligning with the cumulative damage theory of aging, where progressive molecular dysregulation establishes a self-reinforcing feedback loop that exacerbates age-related diseases^45^. In contrast, the traditional FI maintains a relatively constant progression rate irrespective of age or baseline frailty status—consistent with prior studies^46,47^. This divergence underscores fundamental differences in what these indices measure: PFS’s sensitivity stems from its ability to detect subclinical biological processes driving frailty pathogenesis. By capturing dynamic molecular responses to age-related stressors^48^, it identifies early, preclinical frailty manifestations that precede overt functional decline. In contrast, the FI relies on clinical deficits and therefore exhibits limited sensitivity during early aging stages when functional impairment remains subclinical. Its progression rate may be further constrained by ceiling effects in advanced frailty cases^49^, selective survival bias favoring individuals with slower progression trajectories, and the increased likelihood of clinical interventions in severely frail individuals that may attenuate observable functional decline. This dissociation implies that molecular frailty manifests earlier than clinical frailty, but the predictive value of PFS progression for adverse health outcomes and the mechanisms underlying its accelerated trajectory require further investigation.

Our study identifies two critical biological windows for frailty intervention. Unlike previous reports of proteomic change crests at 30-40 years in general aging (e.g., Lehallier et al. reported crests at 34, 60, and 78 years^50^), we found that frailty-associated proteomic changes peak later—first in the late fourth decade (∼50 years) and then consistently at ∼60 years. This delayed onset (peaking at 50 vs. 30-40 years) may be attributed to healthier baseline status of the UKB participants^51^. Moreover, this discrepancy suggests that frailty may represent a distinct physiological process with its own temporal trajectory, rather than simply reflecting accelerated aging. Frailty preventive strategies may need different timing than general anti-aging interventions. Notably, the proteins of these two peaks exhibited stage-specific pathway enrichments, i.e., metabolic and signaling pathways vs. immune-related pathways. This finding suggests that midlife may represent a critical phase for developing metabolic dysregulation that predisposes to later frailty, whereas later life exhibits a transition from metabolic to inflammatory frailty mechanisms^50^. The identification of these two critical windows enables early, pathway-specific interventions targeting distinct biological phases of frailty, prior to clinical onset. Note that, however, these conclusions are based on cross-sectional data and thus should be interpreted with caution despite the large sample size. Longitudinal studies will be essential to confirm the temporal dynamics of these proteomic changes. Future research should also aim to elucidate causal biomarkers and mechanistic drivers of frailty progression to guide targeted therapeutic strategies.

Some limitations in this study should be noted. First, while we characterized plasma proteomic signatures of frailty, its association with tissue-specific protein expression remains uncertain given known inter-tissue variability^52^. Although we employed an established pathway analysis method to eliminate redundant pathways and identified novel functional modules, the plasma-based assay inherently over-represents secreted proteins (particularly immune-related molecules) while under-representing other biological processes, potentially obscuring some molecular mechanisms. Second, external validation of PFS is lacking due to limited availability of large-scale cohorts with comprehensive frailty assessments and Olink large panel proteomic data. Nevertheless, the robustness of our findings is supported by the use of a ten-fold iterative scheme, cross-validation, and sensitivity analyses. Third, while we accounted for numerous modifiable risk factors, certain factors may have been unintentionally neglected. Residual confounding may persist due to collinearities among these factors. Meanwhile, the cross-sectional design precludes causal inference. Fourth, the majority of participants in the UKB were White British and tended to be healthier and wealthier^51^. Further proteomic studies in large-scale non-European ancestral cohorts are needed. Finally, to avoid sample overlap in MR analyses, we utilized FinnGen’s plasma proteome GWAS data generated using the Somascan platform. While moderate technical concordance was observed between SomaScan and Olink platforms, the former identified fewer pQTLs and genomic associations^53^, suggesting these MR results require cautious interpretation and further validation.

In summary, this study delineates the most comprehensive plasma proteomic landscape of frailty in the largest sample to date, identifying novel functional modules implicated in frailty pathogenesis and highlighting promising biomarkers driving its progression. Notably, we pinpointed 50 and 63 years as critical time points for targeted interventions to mitigate frailty progression during aging. These findings bridge key knowledge gaps in understanding the molecular mechanisms of frailty, offering significant translational potential for developing robust biomarkers and personalized therapeutic strategies to promote healthy aging. More importantly, we introduce PFS, a novel proteomic-based frailty measure that demonstrates superior predictive capacity for incident diseases, broad responsiveness to diverse modifiable risk factors, and high sensitivity to preclinical molecular dynamics across aging. As a plasma-based assay derived from routine blood sampling, PFS exhibits advantages of objective quantification and high accessibility. Although current proteomic profiling requires specialized platforms, these technologies are increasingly accessible in research and emerging clinical settings. With continued advancements in scalability and cost-efficiency, PFS hold promise for integration into future clinical workflows and for advancing precision medicine in aging and frailty research.

## 4. Methods

### 4.1 Study participants

The UKB is a prospective, population-based cohort comprising over 500,000 participants aged 40-69 years at baseline, recruited between 2006 and 2010 through 22 assessment centers across the UK. Data were collected via touch-screen questionnaires, physical measurements, biological samples, and linked medical and death register records. Detailed study design and methodology have been reported elsewhere^54^.

As an independent external validation cohort, the TwinGene study, a population-based subcohort of the Swedish Twin Registry (STR), recruited Swedish twins born between 1911 and 1958 who had previously participated in the Screening Across the Lifespan Twin questionnaire (SALT)^55^. Between 2004 and 2008, a total of 12,614 twins were recruited and underwent a basic health checkup, completed a self-reported questionnaire, and provided blood samples.

### 4.2 Plasma proteomic measurements

The UKB Proteomics Project (UKB-PPP) consortium performed proteomic profiling on blood plasma samples collected from 53,026 participants at baseline, using Olink Explore™ Proximity Extension Assay and next-generation sequencing between April 2021 and February 2022. Stringent quality control (refer to <u>biobank.ndph.ox.ac.uk/ukb/ukb/docs/PPP_Phase_1_QC_dataset_companion_doc.pdf</u> for detailed information) was implemented to enable high-throughput quantification of 2,923 proteins across eight panels: cardiometabolic (I and II), inflammation (I and II), neurology (I and II), and oncology (I and II). The final assay read-out was represented as Normalized Protein eXpression (NPX), which was log-transformed and standardized to ensure comparability across samples and batches and accuracy of measuring low-abundance proteins; higher NPX values indicated higher protein expression. Further detailed protocols for sample handling, assay validation, data processing and quality control procedures were provided in previous publications^14,56^. In this study, we retained 2,911 proteins with ≤ 20% missing values. After excluding participants with missing data on > 80% proteins, > 10 frailty items, or covariate information, 50,506 participants were included in the primary analyses. Missing values of proteins were imputed using the k-nearest neighbors (KNN) method.

In the TwinGene cohort, circulating proteins were measured in serum using two different proteomics methods. Olink’s oncology I and II panels^57^ were utilized for indirect protein level measurement in a subgroup, while SWATH^58^ mass spectrometry directly measured protein levels across the complete cohort. In this study, we selected 5,446 participants with available data on 297 proteins overlapping with those significantly associated with FI in the UKB, FI information, along with corresponding FI scores and relevant covariates, for replication analyses.

### 4.3 Frailty assessment

Frailty was assessed using the FI, following the methods proposed by Rockwood et al^4^. In the UKB, the FI was constructed using 49 self-reported items across sensory, cranial, mental well-being, infirmity, cardiometabolic, respiratory, musculoskeletal, immunological, cancer, pain, and gastrointestinal conditions (***Table S46***)^59^. These variables reflect both physiological and mental health, including symptoms, disabilities, and diagnosed diseases. The FI was calculated by dividing the number of deficits present in a participant by the total number of possible deficits, yielding a continuous FI score ranging from 0 to 1. Participants with missing data on more than 10 FI items were excluded. Following validated thresholds^60^, participants were classified as non-frail (FI score < 0.21) and frail (FI score ≥ 0.21). In the TwinGene cohort, the FI was derived from 44 comparable self-reported health items and subsequently calculated using the same method^61^.

### 4.4 Ascertainment of health-related outcomes

Health outcomes were ascertained by linking UKB participants to the National Health Service (NHS) electronic health records using the International Classification of Diseases (ICD)-10 codes. Following the methodology outlined by Deng et al.^62^, we utilized the FinnGen disease endpoints code (https://www.finngen.fi/en/researchers/clinical-endpoints) and adhered to FinnGen’s quality control guidelines. This process included pre-defined conditions based on sex and age, as well as exclusions for specific diseases in the control groups. Notably, several diseases originally coded with two decimal places were rounded to one decimal to align with the ICD-10 code format used by UKB. Incident cases were defined as those occurring after baseline assessment, with follow-up time censored at the earliest occurrence of disease diagnosis, death, loss to follow-up, or end of follow-up (October 31, 2022). A total of 655 incident disease endpoints were included in this analysis, with participants diagnosed before baseline being excluded. Detailed quality control criteria and endpoint specifications were shown in ***Table S31***.

Dates and causes of death were ascertained by linking to national death registries. The all-cause and cause-specific mortality (i.e., cancer, CVD, respiratory disease, neurodegenerative disease, digestive disease, and other causes) were classified according to ICD-10 codes (***Table S47***).^63^ Follow-up time was defined as the time from the baseline assessment date to the earliest of the following events: date of death, date of loss to follow-up, or the end of follow-up (December 31, 2022).

### 4.5 Modifiable factors

We considered a total of 99 potentially modifiable factors derived from the UKB baseline survey^64^. Detailed information on the processing of these factors is provided in ***Table S48***. These factors were categorized into six categories: local environment (e.g., greenspace percentage, buffer 1000m), psychosocial (e.g., nervous feelings), socioeconomic status (SES) (e.g., TDI), early life and sexual health (e.g., breastfed as a baby), physical measures (e.g., handgrip strength), and lifestyle factors (e.g., healthy diet).

### 4.6 Age-related chronic diseases, metabolic indicators, and inflammatory markers

We tested associations between the proteins of two key crest and health-related traits, including age-related chronic diseases, metabolic indicators, and systemic inflammatory markers. Chronic diseases included CVD, cancer, dementia, diabetes, chronic kidney disease, liver diseases, respiratory disease, osteoarthritis, depression, Parkinson’s disease, multiple sclerosis, and schizophrenia. These diseases were selected based on lifelong contribution to substantial health burden or brain-associated illness burden (i.e., depression and schizophrenia) in older adults^65^. Metabolic indicators included TC, TG, and high-density lipoprotein cholesterol. Inflammatory markers included baseline counts of neutrophils, monocytes, platelets, and lymphocytes, and level of C-reactive protein. We further calculated four ratios based on peripheral blood cell counts including NLR (neutrophils/lymphocytes), PLR (platelets/lymphocytes), LMR (lymphocytes/monocytes), SII (neutrophils × platelets/lymphocytes), and SIRI (neutrophils × monocytes/lymphocytes).

### 4.7 Covariates

Demographic variables included age, sex (female or male), ethnicity (White or non-White), and educational level (high, intermediate, or low). Lifestyle factors included smoking status (never, previous, or current smoker), alcohol intake frequency (never or special occasions only, 1-3 times monthly, 1-4 times weekly, or daily/almost daily), regular exercise (yes/no), and healthy diet (yes/no). Anthropometric data included body mass index (BMI, kg/m²), calculated as weight/height^2^. The first 20 genetic principal components (PCs) derived from genome-wide genotyping data were included to account for population stratification and genetic background differences.

### 4.8 GWAS summary statistics of frailty

We obtained GWAS summary statistics for frailty (measured using FI) via the European Bioinformatics Institute (EBI) database (accession: ebi-a-GCST90020053). This GWAS study included 175,226 individuals of European ancestry, comprising 164,610 participants from the UKB aged 60-70 years and 10,616 participants from the Swedish TwinGene study aged 41-87 years^27^.

### 4.9 Data sources of pQTL and eQTL

The pQTLs are genetic variations associated with protein levels, while expression quantitative trait loci (eQTLs) are linked to gene expression levels. We utilized data from a large-scale GWAS conducted by the deCODE Genetics team, which employed plasma proteomics technology to analyze 4,907 proteins in 35,559 individuals from Iceland. Additionally, we curated eQTL data from the eQTLGen Consortium (https://eqtlgen.org/)^66^, which included 16,989 cis-eQTL genes identified through genetic analysis of 31,684 blood samples from healthy individuals of European ancestry. The pQTL and eQTL datasets used in our analyses are derived from non-overlapping cohorts relative to the FI GWAS dataset.

### 4.10 Statistical analysis

#### 4.10.1 Participants characteristics

We presented the baseline characteristics of the analytic samples overall and by frailty status. Categorical variables are shown as numbers and percentages, and continuous variables are shown as medians with IQRs. Differences between frailty groups were assessed using χ² tests for categorical variables and Kruskal-Wallis tests for continuous variables.

#### 4.10.2 PWAS of frailty

Linear regression models were performed to assess the associations between protein levels (as the predictor) and FI (as the outcome). The β coefficients and corresponding 95% CIs were estimated using two models. Model 1 was adjusted for age, sex, and the first 20 genetic PCs, and Model 2 was additionally adjusted for ethnicity, educational level, TDI, smoking status, alcohol intake frequency, regular exercise, healthy diet, and BMI. The fully adjusted model (Model 2) was used for all primary analyses.

To ensure the robustness of our findings, we conducted several sensitivity analyses: 1) Treating frailty as a dichotomous variable (non-frail vs. frail), with non-frail participants as the reference group; 2) Restricting the analyses to participants with complete data on all FI items; 3) Restricting the analyses to a randomly selected subset to address potential bias from the UKB-PPP sub-cohort composition; 4) Focusing specifically on Caucasians to further mitigate the possibility of population stratification; 5) Incorporating interaction terms between sex or age and protein levels into the models to evaluate sex- and age-related differences. Additionally, subgroup PWAS analyses were performed separately for males and females, as well as for younger (< 60 years) and older (≥ 60 years) individuals, to provide further insights into potential heterogeneity.

For replication, two linear regression models were used to examine the associations between circulating proteins and the FI in the TwinGene cohort. Model 1 was adjusted for age and sex, while Model 2 was additionally adjusted for education years, smoking status, and alcohol consumption in the year prior to blood sampling, and BMI. To address the non-independence of observations within twin pairs and account for the twin structure of the data, cluster-robust standard errors were utilized. To better meet the assumptions of linear regression, the FI was log-transformed (with a 0.1 offset) and multiplied by 10 to improve interpretability.

#### 4.10.3 Pathway enrichment analysis and functional module identification

Pathway enrichment analysis was performed using the clusterProfiler R package, referencing the Gene Ontology (GO), Kyoto Encyclopedia of Genes and Genomes (KEGG), and Reactome databases^67^. Adjusted *P* values were calculated using the Benjamini-Hochberg method, with a significance threshold set at < 0.05. To eliminate redundancy, we calculated the similarity between enriched GO terms and pathways: the ‘Wang’ algorithm from the simplifyEnrichment R package was used for GO terms, while the ‘jaccard’ algorithm was applied to KEGG and Reactome pathways. Community analysis was then conducted using the igraph R package to partition the similarity network into distinct modules based on edge betweenness. Within each module, the pathway or GO term with the smallest adjusted *P* value was selected as the representative one. Then we merged all the remaining pathways from all three databases and repeated the above process by recalculating the similarity using the Jaccard algorithm. Finally, we employed the same approach again to identify overarching functional modules.

#### 4.10.4 The development of PFS model

To develop PFS, we performed ten iterations of LASSO modeling with ten-fold cross-validation using the Caret R package. The model was applied to the log-transformed (FI + 0.1) as the outcome, with 1,339 frailty-associated proteins identified through PWAS as predictors. Participants were split into ten sets, with one set designated as the testing (hold-out) set and the remaining nine as the training set. This process was iterated across all ten combinations to generate ten fitted sparse models and one hold-out set-derived prediction for each participant. Overfitting was controlled through ten-fold iteration, and hyperparameters were optimized using ten-fold cross-validation within each training set. To ensure balanced splits, we confirmed no significant differences in sociodemographic, lifestyle-related, or genetic background characteristics (i.e., the first 20 genetic PCs) among the ten sets using χ² tests for categorical variables and ANOVA for numeric variables (***Table S25***).

Model performance was evaluated using the out-of-sample R^2^ and mean absolute error (MAE), calculated from each corresponding hold-out testing set. Additionally, Pearson’s correlation coefficient (r) between the measured and predicted values were computed across all participants.

#### 4.10.5 The predictive performance of the PFS and FI for health-related outcomes

We then estimated the predictive performance of PFS and FI for diseases risk. First, PFS and FI were used independently for disease-risk prediction. Performance was quantified by Harrell’s C-index using survival R package. Then, four multivariable prediction models with different combinations of PFS and FI and conventional risk factors (age, sex, ethnicity, TDI, smoking status, alcohol intake frequency, BMI, and systolic blood pressure) were built as follows: Model 1 (only conventional risk factors), Model 2 (conventional risk factors + PFS), Model 3 (conventional risk factors + FI) and Model 4 (conventional risk factors + PFS + FI). These models were built using Cox proportional hazards regression for each disease. The differences in Harrell’s C-indexes of these models were compared using the compareC R package. Benjamini-Hochberg-adjusted *P* values < 0.05 were considered statistically significant.

#### 4.10.6 Associations of modifiable factors with PFS and FI

Multivariable linear regression models were used to evaluate the associations between each modifiable factor (independent variable) and PFS or FI (dependent variable). A Bonferroni-corrected significance threshold was applied to identify significant associations (*P* < 0.05/99 = 5.05×10^−4^). In these analyses, continuous variables were standardized (z-normalized), and results were expressed as β coefficients per 1-SD increase in the corresponding factor. Modifiable factors were further categorized into tertiles, with the lowest tertile serving as the reference group. All models were adjusted for age, sex, and ethnicity. Stratified analyses were performed by age group (< 60 and ≥ 60□years) and sex (male and female).

#### 4.10.7 Calculation of PFS (and FI) change rate with revisit proteome data

UKB includes unique follow-up plasma proteomics data for 1,161 participants at their third visits and 1,113 participants at their fourth visits. For longitudinal analyses, we focused on 1,459 proteins with less than 20% missing values across all three visits (first, third, and fourth). We re-trained PFS models using these 1,459 proteins (“feature-reduced” models) in the 50,506 participants at baseline, following the same approach described above.

For analyses of PFS (and FI) change rates, we retained 784 participants with less than 80% missing data in the 1,459 proteins across all three visits and complete visit date information (Field 53). Missing values of proteins were imputed using the KNN method. For each participant at each time point, the PFS prediction was obtained by averaging ten predicted values generated from the baseline LASSO models applied to the processed time-series proteomic data. Change rates of frailty were then calculated based on the following mixed-effects model^68^:

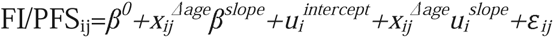

PFS_ij_ (and FI_ij_) represents the predicted PFS (and FI) for participant *i* at visit *j*. The fixed intercept is denoted by β^0^, and 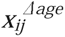 represents the difference in age between visit *j* and the first visit for participant *i*. The fixed effect β*^slope^* captures the average annual change in predicted PFS (and FI). Participant-specific intercepts and slopes are modeled through random effects 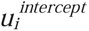 and 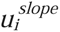, respectively, while *ε_ij_* represents the random error term. All random effects and error terms are assumed to follow normal distributions. The mixed-effects models were performed using the lmer function from the lme4 R package. For each individual *i*, the longitudinal slope was calculated as 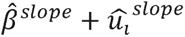 (where the hats indicate the estimated values of the corresponding model parameters), corresponding to the average rate of change in PFS (and FI) during the follow-up period.

We then assessed the correlation between the PFS (and FI) change rate and baseline PFS (and FI) using Spearman correlation coefficient. Additionally, we evaluated the association between the PFS (and FI) change rate and accumulated disease counts using linear regression models, adjusting for the covariates listed above. To validate the robustness of our findings, we also calculated the PFS (and FI) change rate by subtracting the previous round from the subsequent round divided by follow-up time.

#### 4.10.8 DE–SWAN

The DE-SWAN method, implemented using the DEswan R package, was used to quantify nonlinear changes in the frailty-related proteome during aging^50^. Participant ages were rounded, and the analysis was restricted to participants aged 39 to 71 years. A sliding window approach was employed, with a 6-year window comparing molecule levels between groups in parcels of 3□year, then sliding the window in increments of 1□year from young to old age. Differential expression was assessed using the Wilcoxon test, and *P* values were adjusted for multiple comparisons using the Benjamini-Hochberg correction. Covariates including sex, the first 20 genetic PCs, ethnicity, educational level, TDI, smoking status, alcohol intake frequency, regular exercise, healthy diet, and BMI were adjusted in the DE-SWAN analysis. To evaluate the robustness of the results, the algorithm was tested with different q-value thresholds (<0.0001, <0.001, <0.01, and <0.05).

#### 4.10.9 Organ-specific proteins enrichment analyses

Following the methodology by Oh et al.^25^, we mapped the putative organ-specific plasma proteome by defining a protein as organ-specific if its corresponding RNA expression in the Genotype-Tissue Expression (GTEx) database was at least four-fold higher in one organ compared to any others. Fisher’s exact test was used to evaluated whether frailty-associated proteins were significantly enriched with specific organs.

#### 4.10.10 Proteome-wide MR analyses

The pQTLs from the aforementioned proteomic study were used for the selection of genetic instruments. The following criteria were applied for selecting instruments and proteins: (i) SNPs associated with any protein were selected (*P* < 5×10^−8^); (ii) SNPs and proteins within the Major Histocompatibility Complex (MHC) region (chr6:25.5-34.0Mb) were excluded due to their complex LD structure; (iii) LD clumping was performed to identify independent pQTLs for each protein (r² < 0.001); and (iv) the R² and F-statistic were used to estimate the strength of genetic instruments, where R² was the proportion of the variability of protein levels explained by each genetic instrument, and SNPs with an F-statistic less than 10 were excluded. The TwoSampleMR R package was employed to perform MR analysis. The Wald ratio method was used to obtain MR effect estimates for proteins with only one instrument. The IVW method was used to obtain MR effect estimates for proteins with more than one instrument. A heterogeneity test was performed to assess the heterogeneity of the genetic instruments based on the Q statistic. We also conducted additional analyses, including simple mode, weighted mode, weighted median, and MR-Egger, to account for horizontal pleiotropy. MR-Egger results were used only when the intercept indicated the presence of horizontal pleiotropy. A *P* value < 0.05 was considered statistically significant.

#### 4.10.11 PheMR analyses

PheMR is a robust tool to evaluate the potential causal associations between selected proteins and a broad spectrum of phenotypes. In this study, we used outcome data from FinnGen release R5 (https://r5.risteys.finngen.fi/), which includes 2,803 phenotypes. SNPs identified in the pQTL analysis were employed as IVs in the PheMR analysis to evaluate their potential causal effects on these 2,803 phenotypes. The IVW method was used to estimate the potential causal effects. A *P* value < 0.05 was considered statistically significant.

#### 4.10.12 Colocalization analyses

Colocalization analysis used five posterior probability hypotheses: i) no shared causal variants for either trait (H0); ii) causal variant for gene expression only (H1); iii) causal variant for disease risk only (H2); iv) distinct causal variants for each trait (H3); and v) a shared causal variant for both traits (H4). The colocalization window was set at ± 250 kb. This approach was designed to bolster evidence on associations between genetic targets and phenotypes (or disease), facilitating the identification of causal relationships while reducing the risk of confounding variables. The colocalization analysis was performed using the coloc R package^69^, with a threshold of PPH3 + PPH4 > 0.8. This threshold was applied to determine whether shared genetic effects existed between the targets and diseases, providing a solid foundation for subsequent research and therapeutic advancements^70^.

#### 4.10.13 Single cell-type expression analysis

The cell type-specific expression of target genes, which may have a potential causal effect on frailty at the plasma protein levels, was further evaluated using single-cell RNA-seq data of PBMC from frail older individuals (age > 65 years, FI > 0.2) and non-frail older individuals (age > 65 years, FI ≤ 0.2) from the Gene Expression Omnibus (GEO) dataset by Luo et al.^30^ Using the Seurat package, we first conducted data preprocessing and transformation based on the raw single-cell RNA-seq data. Cell type annotations were performed using the SingleR package. Differential expression analysis was then performed using the Wilcoxon Rank Sum test to compare gene expression levels across different cell types between frail and non-frail individuals. The genes with a *P* value < 0.05 were identified as differentially expressed genes.

## Authors’ contributions

Z.Y.L. designed and supervised the study. X.J., W.G., H.H., and J.Z. analyzed the data. X.J. and Y.Z. drafted the manuscript. X.J., W.G., H.H., and Z.Y.L. interpreted the results. All authors revised the manuscript. Z.Y.L. took responsibility for the content of the article. All authors have read and approved the submitted version of the manuscript.

## Data Availability Statement

The data supporting this study’s findings are openly available in UK Biobank at [https://www.ukbiobank.ac.uk/], reference number [61856]. We acknowledge the Swedish Twin Registry for access to data. The Swedish Twin Registry constitutes a national research infrastructure open to projects initiated from both academic and industry-based researchers.

## Ethics approval and consent to participate

UK Biobank has approval from the North West Multi-Centre Research Ethics Committee as a Research Tissue Bank approval in 2011 and is renewed every 5 years, which allowed researchers to use data from UK Biobank without an additional ethical clearance. All participants have provided signed informed consent. The use of data from the Swedish Twin Registry was approved by the Swedish Ethical Review Authority (Dnr 2022-06634-01).

